# The impact of localization and registration accuracy on estimates of deep brain stimulation electrode position in stereotactic space

**DOI:** 10.1101/2024.03.01.24303286

**Authors:** Mohamad Abbass, Alaa Taha, Greydon Gilmore, Brendan Santyr, Alan Chalil, Mandar Jog, Keith MacDougall, Andrew G. Parrent, Terry M. Peters, Jonathan C. Lau

## Abstract

Effects of deep brain stimulation (DBS) depend on millimetric accuracy and are commonly studied across populations by registering patient scans to a stereotactic space. Multiple factors contribute to estimates of electrode position, but the millimetric contributions of these factors remains poorly quantified. We previously validated 32 anatomical fiducials (AFIDs) to measure AFID registration error (AFRE), which can capture focal misregistration not observed using volume-based methods. To this end, we used the AFIDs framework to examine the effects of misregistration on electrode position in stereotactic space, leveraging a retrospective series of patients who underwent subthalamic nucleus (STN) DBS. Raters independently localized DBS electrodes and AFIDs on patient scans, which were non-linearly registered to a common stereotactic (MNI) space. AFIDs provided intuitive measures of registration accuracy, with AFREs ranging from 1.49 mm to 6.85 mm across brain regions. Subcortical AFIDs in proximity to the DBS target (STN) had AFREs that spatially covaried, suggesting consistent spatial patterns of misregistration to stereotactic space. These identified spatial patterns account for 28% of the variance in electrode position along the axis of maximum variance, corresponding to a median of 0.64 mm (range of 0.05 to 2.05 mm). The AFIDs framework provides millimetric estimates of registration accuracy in DBS, while allowing the uncoupling of registration-related factors from other sources of variance in electrode position. Furthermore, they can be employed for estimating registration-related variance in population studies, for quality control, and to provide a basis for comparison as well as optimization of registration parameters and software.

## 1 Introduction

Stereotactic mapping involves affixing an external 3-dimensional (3D) coordinate system (i.e., Cartesian coordinates, with x, y, and z axes) for the identification and treatment of specific brain structures with millimetric accuracy (Horsley & Clarke, 1908; Leksell, 1949; Spiegel et al., 1947). Deep brain stimulation (DBS) employs stereotactic principles for the precise implantation of electrodes in the brain from which focal electrical therapy is applied to treat different disorders. Accurate implantation of DBS electrodes is a critical step, as millimetric deviations have been shown to result in suboptimal clinical outcomes (Li et al., 2016). Postoperatively, imaging including computed tomography (CT) and magnetic resonance imaging (MRI) is used for electrode localization. To study the effects of DBS across a population, individual patient scans are commonly registered to a stereotactic space, facilitating group-level statistics (Horn, 2019; Horn et al., 2017; Zhang et al., 2021). Numerous registration methods have been reported, employing different combinations of linear and nonlinear transformations as well as associated parameters (Evans et al., 1993; V. S. Fonov et al., 2009; Schönecker et al., 2009).

Reliable and accurate identification of DBS electrodes in stereotactic space facilitates population inferences that can guide clinical practice (Barow et al., 2014; Jeon et al., 2022). Electrode position in stereotactic space may vary for multiple reasons including: (1) application accuracy, referring to the “true” variance in position from surgical implantation of the DBS electrode (Cardinale et al., 2017; Henderson et al., 2004); (2) the variability of electrode localization on the post-operative scan; (3) the accuracy of co-registration between the preoperative and post-operative scans; and (4) the accuracy of registration between the patient scan and stereotactic space. Inter-rater electrode localization distance (2) has been investigated in one study (Lofredi et al., 2022) where a mean inter-rater distance of 0.57 ± 0.2 millimeters (mm) was found. The variability in electrode positions following different co-registration parameters (3) has also been investigated, with errors ranging from 0.57 to 1.17 mm (Bower et al., 2023; Engelhardt et al., 2018; O’Gorman et al., 2009). Registration accuracy to a stereotactic space (4) remains poorly quantified.

For neuroimaging applications, the quality of registration to stereotactic space has been commonly assessed with voxel-overlap measures which are based on ratios of spatial correspondence between homologous regions of interest (ROIs), most commonly subcortical structures such as the thalamus or basal ganglia (Ewert et al., 2019; Fonov et al., 2011; Vogel et al., 2020). Voxel-overlap measures are straightforward to obtain from common neuroimaging workflows but are relatively coarse and do not capture focal misregistration (Rohlfing, 2012). Anatomically placed points (also referred to as fiducials or landmarks) can also be used to quantify registration accuracy measured as the millimetric distance between transformed points and their homologous counterpart in stereotactic space (Abbass et al., 2022; Lau et al., 2019; Schönecker et al., 2009). This motivated our group to identify and validate a protocol for the placement of 32 anatomical fiducials (AFIDs) providing a point-based sampling of multiple brain structures with an emphasis on the deep brain (Lau et al., 2019). AFIDs can be localized to within 2 mm by both novice and experienced human raters across research and clinical grade MRI scans (Lau et al., 2019; Abbass et al., 2022; Taha et al., 2023). The AFIDs framework can capture subtle registration errors not observed using voxel-overlap methods (Lau et al., 2019). Additionally, AFIDs can be placed efficiently and incorporated into workflows, requiring users to place a single point for each anatomical region of interest. Finally, point-based registration error can be intuitively interpreted as a vector in space with components of magnitude and direction that can be helpful for understanding the spatial bias of registration methods.

To this end, we sought to investigate the impact of registration accuracy on estimates of DBS electrode position in stereotactic space. For this purpose, we leverage a dataset of patients implanted with DBS electrodes for Parkinson’s disease (PD). First, we replicated the results of Lofredi et al., (2022) obtaining sub-millimetric inter-rater electrode localization distance. We then quantified registration accuracy at various brain locations and found that registration errors across a subset of subcortical AFIDs significantly covaried, suggesting consistent spatial patterns of misregistration. Finally, we demonstrated that these identified spatial patterns explain a significant amount of variance in electrode location in stereotactic space. Overall, the AFIDs framework provides a simple and intuitive method to obtain registration accuracy and capture the variance in the position of DBS electrodes as it relates to registration to stereotactic space.

## 2 Materials and Methods

### 2.1 Patient Selection

We conducted a retrospective analysis of patients who underwent bilateral subthalamic nucleus (STN) electrode placement for PD at our center between 2009 and 2018. All subjects underwent a pre-operative MRI scan that served as the basis for surgical planning as well as a reference scan for image co-registration (described below). All clinical data were obtained from the electronic health records; any missing data were obtained from paper charts. The study was approved by the Human Subject Research Ethics Board (HSREB) office (REB# 109045).

### 2.2 Data Acquisition, Processing, and Annotation

Prior to surgery, a gadolinium-enhanced volumetric T1-weighted (T1w) MRI scan was acquired (echo time = 1.5 ms, inversion time = 300 ms, flip angle = 20°, receiver bandwidth = 22.73 kHz, field of view = 26 cm × 26 cm, matrix size = 256 × 256, slice thickness = 1.4 mm, resolution = 1.25 × 1.25 × 1.50 mm; Signa, 1.5 T, General Electric, Milwaukee, Wisconsin, USA). Once surgery was complete, a postoperative non-contrast MRI or CT scan was acquired for the purpose of localizing the DBS electrode.

Using default Lead-DBS (v.2.3.2) parameters (Horn & Kühn, 2015), each subject’s postoperative CT or MRI was linearly registered to the preoperative MRI as implemented in Advanced Normalization Tools (ANTs; Avants et al., 2008; “http://stnava.github.io/ANTs/”). The reference volume was then nonlinearly registered to the MNI152NLin2009bAsym (MNI) space (Fedorov et al., 2012) using the SyN registration approach in ANTs. Nonlinear deformation into MNI space was achieved in five stages: following two linear (rigid followed by affine) steps, a nonlinear SyN registration stage was followed by two nonlinear SyN registrations that consecutively focused on the area of interest as defined by subcortical masks (Schönecker et al., 2009), which is recommended for DBS studies. We independently registered volumes to MNI space using fMRIPrep (v.1.5.4) with default parameters (Esteban et al., 2019). A summary of registration parameters used in Lead-DBS and fMRIPrep is provided in **Table S1**.

Four expert raters (see **Table S2** for demographics) were recruited to localize DBS electrodes. Raters were randomly paired (group 1: raters A/B, group 2: raters C/D) for sufficient rater sampling across the dataset. For each patient, the Lead-DBS workflow was independently run by one rater from each group. Electrodes were semi-automatically localized using Lead-DBS, and manually adjusted by each rater. Raters subsequently localized the anterior commissure (AC) and posterior commissure (PC) on the patient’s preoperative MRI, as defined by clinical practice (Fedorov et al., 2012; Horn et al., 2017). In addition, ground truth AC and PC were defined by consensus among all raters in MNI space with the following coordinates (x, y, z): AC (−0.24, 1.88, −4.75) and PC (−0.06, −24.68, −2.36), visualized in **Figure 1a**. Finally, we leverage previously validated and openly released AFID annotations in a subset of this study’s cohort (Abbass et al., 2022; Taha et al., 2023). Briefly, five raters independently localized 32 AFIDs on subject preoperative MRI and on the MNI template.

**Figure 1.**
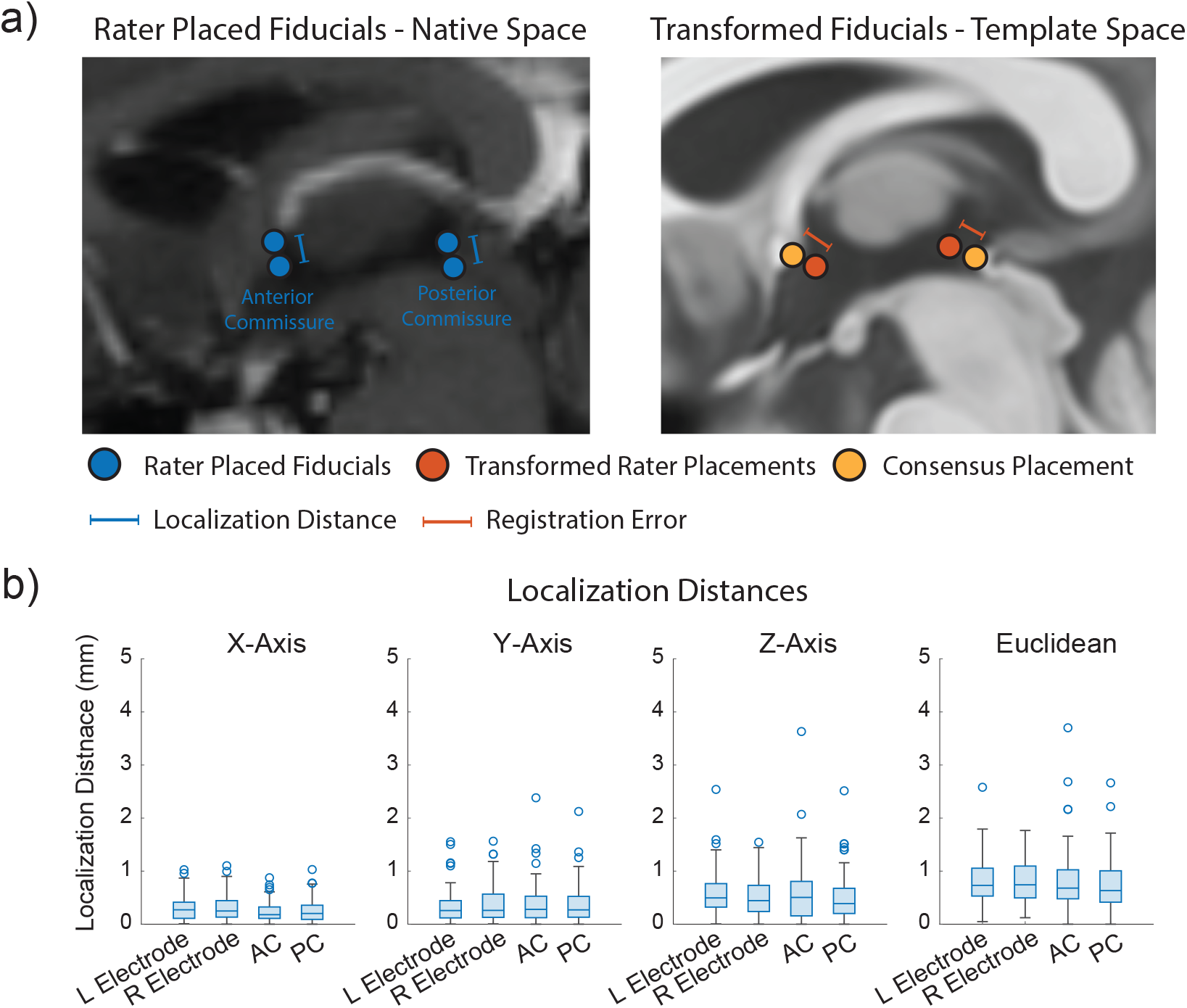
Summary of methodology with localization accuracy of deep brain stimulation electrodes and anatomical fiducials. (a) Schematic depicting localization distance and registration error. Localization distance is shown as the distance between rater placed coordinates within a subject’s native space, and registration error is shown as the displacement between raters’ transformed coordinates (Transformed Rater) to rater placed coordinates (Consensus) in MNI space. (b) Localization distances of DBS electrodes, intraventricular anterior commissure (AC) and posterior commissure (PC) obtained in each axis, and the Euclidean localization distance. There were no significant differences in localization distance between electrode, AC, and PC LEs in all axes (Wilcoxon Sign-Rank Test (n=89), alpha = 0.05/6 in each axis).

### 2.3 AFID Localization Error and Electrode Localization Distance

In prior work, we defined the **AFID Localization Error (AFLE)** to describe the Euclidean distance between rater AFID localizations and a curated ground truth (Lau et al., 2019; Abbass et al., 2022; Taha et al., 2023). This ground truth required that two key elements be satisfied: (1) a validated open-access protocol describing the predefined positions of AFIDs and (2) averaging placements from three or more independent rater annotations to curate consensus coordinates.

In the present study, some of the measurements performed, including electrode position, were not anatomically “predefined”, and were only performed by two raters per subject, which would not satisfy the elements of our “localization error” definition, and could be misleading. In this scenario, we use the term inter-rater Localization Distance (LD), calculated as the Euclidean distance between the localizations of raters (i.e., pairwise distance), a definition which is consistent with what is used in prior work (Lofredi et al. 2023). We compared LDs of DBS electrodes, AC, and PC using a Wilcoxon signed-rank test, and used a Bonferroni correction for multiple comparisons in each axis. Additionally, given that electrodes were localized on post-operative CT and MRI scans, we compared LDs between these modalities using a Wilcoxon rank sum test. For completeness, we also summarize AFLE in the subset with AFIDs annotations.

### 2.4 Registration Accuracy

Each rater’s manual AC and PC annotations were transformed to MNI space. We calculated the displacement between each rater’s transformed AC and PC to the homologous ground truth MNI placements across the x, y, and z axes, and the Euclidean distance, a metric of registration accuracy previously defined as the **AFID Registration Error (AFRE)** in Lau et al., (2019). **Figure 1a** provides an illustration summarizing the calculation of inter-rater LD and AFRE. We compared inter-rater LDs and AFREs obtained using a Wilcoxon signed-rank test. AFRE was computed in a similar manner for the subset of patients who had the AFIDs protocol. Specifically, we calculated the mean AFID coordinates across raters for each subject and applied the transforms obtained from Lead-DBS v2.3.2 to MNI space to compute AFRE. We performed the same analysis using the transforms obtained from fMRIPrep v1.5.4.

AFREs were compared using a Wilcoxon signed-rank test with Bonferroni correction to the number of AFIDs. A p-value less than 0.05, but not meeting correction thresholds of statistical significance were also highlighted. Additionally, we further expanded our analysis to accommodate for new software releases since this initial project was started. We directly compared AFREs obtained with the aforementioned software versions to registrations completed in Lead DBS v3.1.0 and fMRIPrep v21.0.1 using a Wilcoxon signed-rank test.

### 2.5 Correlating AFRE and Electrode Position

AFREs provide vectors of registration error in both magnitude and direction at AFID locations rather than specific points of interest, such as an electrode contact. Across subjects, different AFREs may covary if there are consistent patterns of misregistration. Additionally, variance in electrode tip position in stereotactic space may be explained by AFREs. To this end, we investigated whether AFREs obtained at AC and PC were correlated with DBS electrode positions in stereotactic space. We centered the electrode tip locations by subtracting each patient electrode tip coordinate from the mean coordinate of all electrode tips. The displacement of each subject’s electrode tip from the mean electrode tip location was correlated with the AFRE obtained in AC and PC, and for each x, y, and z axis. We performed a simple linear regression to measure the displacement in electrode position explained by AFRE in each axis and used a Bonferroni correction for multiple comparisons for each AFID. We also explored other potential explanatory variables, including age, disease duration, sex, rater pair, modality (post-operative CT or MRI used), electrode side and implantation order. These variables were individually tested using univariate non-parametric tests, using a Wilcoxon rank sum test for binary variables (sex, rater pair, modality, side and implantation order) and a Spearman’s rank correlation for continuous variables (age and disease duration). A Bonferroni correction for multiple comparisons for each explanatory variable. A multivariate linear regression was also used including all variables and AFREs at AC and PC.

We further analyzed a subset of AFIDs which were found to explain a significant amount of variance in the electrode tip position. We first performed principal components analysis (PCA) on individual AFREs obtaining three orthogonal unit vectors (principal components; PrCs), representing the independent axes explaining the variance of AFREs. We performed this same analysis on the displacement of the electrode tip. To explore the relationship between AFREs, we computed a correlation matrix by performing simple linear regressions between all pairwise AFREs across the x, y, and z axes. The same analysis was performed with AFREs projected along their PrCs. Finally, we obtained high-dimensional axes explaining consistent variance across these AFIDs by performing PCA on all axes of the AFIDs used (3 axes x 4 AFIDs, or 12 features). We examined whether AFREs projected onto these PrCs were correlated with the electrode tip positions using a simple linear regression.

## 3 Results

### 3.1 Patient Demographics

Data from 89 patients implanted with bilateral STN DBS successfully underwent the Lead-DBS protocol independently by two rater groups for electrode localization. These patients had a mean age of 60.54 ± 6.12 years, a mean disease duration of 11.01 ± 4.21 years, and 31 were female (31.46%). Of these patients, 24 had the AFIDs protocol previously completed (Abbass et al., 2022). See **Table S3** for a summary of the demographic variables of these patient populations.

### 3.2 AFLE and Inter-Rater LD

We first sought to investigate inter-rater LD of DBS electrodes, the anterior commissure (AC), and the posterior commissure (PC) by calculating the absolute difference between the two rater placements in each axis and across all axes (Euclidean error). **Figure 1b** summarizes the LD for each electrode tip, AC, and PC (see **Table S4** for a complete summary of LDs). The median interrater LD (with interquartile range; IQR) was 0.73 mm (0.53-1.06 mm) for the right electrode tip, 0.74 mm (0.50-1.10mm) for the left electrode tip, 0.68 mm (0.48-1.03 mm) for AC, and 0.63 mm (0.41-1.01 mm) for PC. A Wilcoxon signed-rank test revealed no significant differences between any LDs in all axes. Exploring the modality used for post-operative imaging, LD for all electrode tips was 0.57 mm (0.32-0.73 mm) using CT (n=19) and 0.83 mm (0.57-1.14 mm) using MRI (n=70), which was statistically significant (p<0.001, Wilcoxon rank sum test).

We expanded our analysis to a subset of 24 patients with ground truth AFID annotations, derived as the average placement of five human raters who independently annotated scans. The median AFLE ranged from 0.56 mm (0.40-0.69 mm) for AFID02 (PC) to 2.25 mm (1.49-2.75 mm) for AFID25 (right inferior anteromedial temporal horn). See Abbass et al. (2022) for more details.

### 3.3 Registration Accuracy

So far, we have described our calculation of LD, which represents a measure of electrode localization reliability; however, registration to stereotactic space introduces additional error. To capture a fiducial-based measure of this error at the AC and PC locations, rater-placed coordinates were transformed to MNI space. AFREs are visualized as a 3D point cloud centered around the consensus placement (**Figures 2a/b**) and summarized in **Table S5**. The median Euclidean AFRE with IQR was 1.39 mm (1.05-2.38 mm) for AC (**Figure 2c**) and 1.42 mm (1.00-2.10 mm) for PC (**Figure 2d**). Compared to LD at AC and PC, AFREs at these locations were significantly greater in the y and z axes (Wilcoxon signed-rank Test; **Figure 2c/d**). The same AFREs at AC and PC were independently obtained across the four raters (**Figure S1**).

**Figure 2.**
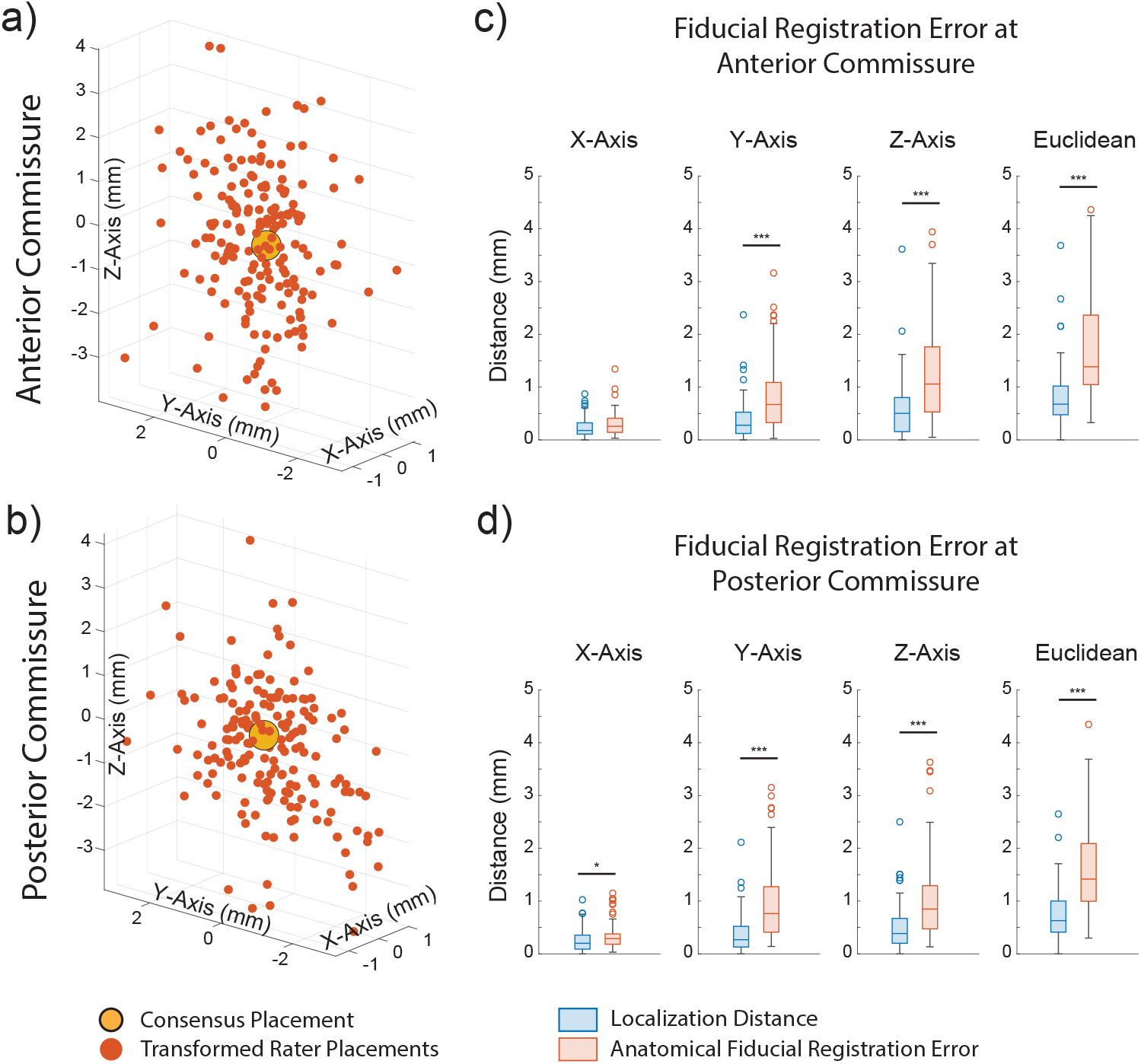
Anatomical fiducial registration errors at anterior and posterior commissures in MNI space. (a-b) 3D scatterplots represent transformed rater placements (red) of anterior commissure (AC) and posterior commissure (PC), and the consensus placement (yellow). The displacement of each transformed rater placement from the consensus placement represents its anatomical fiducial registration error (AFRE). (c-d) Boxplots comparing AFRE to localization distance (LD) at AC and PC in all axes (Wilcoxon Sign-Rank Test (n=89), * p<0.05, *** p<0.001). AFRE was significantly greater than LD at AC and PC in the y and z axes (p<0.001).

We next expanded our analysis of AFRE by using the entire set of previously defined and validated AFIDs (Abbass et al., 2022; Lau et al., 2019). **Figure 3** summarizes Euclidean AFREs obtained for each AFID and illustrates 3D point-clouds of AFREs. For each subject, the mean AFRE across all AFIDs was calculated as a global measure of AFRE. The median (IQR) global AFRE was 3.09 mm (2.80-3.22 mm). AFRE was not uniform across AFIDs, with centrally located subcortical AFIDs having lower AFREs. The lowest AFREs were obtained at AFID03 (infracollicular sulcus, ICS) and AFID01 (AC) with AFREs of 1.49 mm (0.96-2.25 mm) and 1.59 mm (0.69-2.31 mm) respectively. The highest AFREs were obtained around the ventricles, the highest of which being AFID29 and AFID30 (right and left ventral occipital horn), with AFREs of 6.85 mm (4.47-7.90 mm) and 6.61 mm (5.70-8.69 mm) respectively.

**Figure 3.**
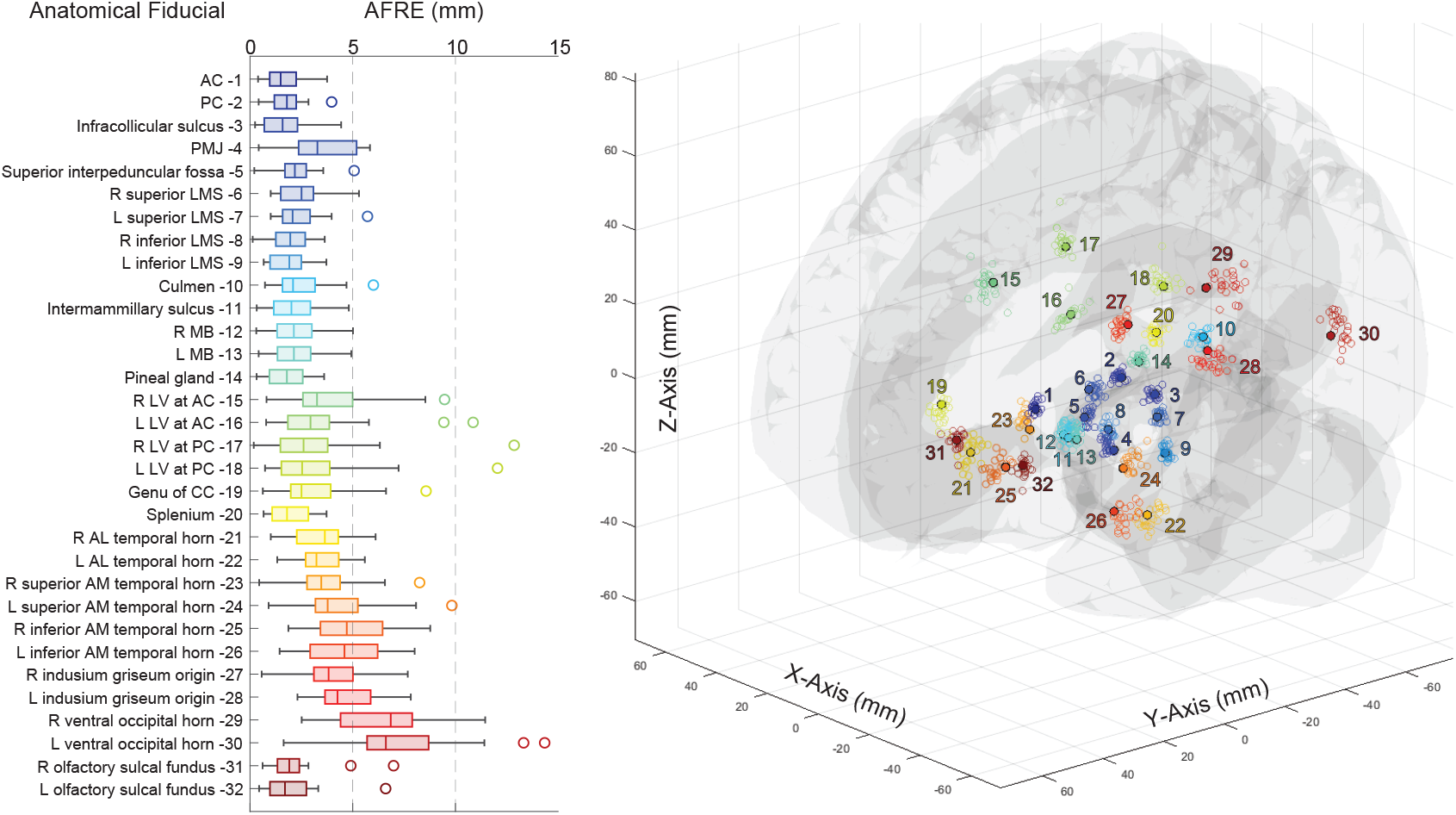
Anatomical fiducial registration errors for the complete set of 32 anatomical fiducials. Boxplot of Euclidean fiducial registration errors (AFREs) calculated for 32 anatomical fiducials (AFIDs; left) and 3D scatterplot of transformed rater AFID placements (n=24; open circles) with consensus AFID placements (filled circles) in MNI space (right). Euclidean AFRE ranged from a median (with IQR) of 1.49 mm (0.96-2.25 mm) for AFID 03 to 6.85 mm (4.47-7.90 mm) for AFID 29. Subcortical and midline AFIDs had the lowest AFREs, while peripherally located and periventricular AFIDs had the highest AFREs.

In previous work, we explored AFREs obtained from these same subjects following an automated non-linear registration to MNI space using fMRIPrep 1.5.4 (Abbass et al., 2022; Esteban et al., 2019). **Figure S2** summarizes the Euclidean AFREs obtained for all AFIDs using both the Lead-DBS and fMRIPrep pipelines (for the specific versions outlined). Overall, the global AFRE was higher using Lead-DBS (3.09 mm, 2.80-3.22 mm) when compared to fMRIPrep (2.75 mm, 2.46-3.01 mm; p = 0.002). This difference was not homogenous across AFIDs, and even with a stringent Bonferroni correction, 2 AFIDs had lower AFREs when using Lead-DBS: AFID14 (pineal gland, PG), and AFID20 (splenium). However, 6 AFIDs had higher AFREs using Lead-DBS: AFID01 (AC), AFID04 (pontomedullary junction, PMJ), AFID05 (superior interpeduncular fossa), AFID21 (right anterolateral temporal horn), AFID25 (right inferior anteromedial temporal horn), and AFID26 (left inferior anteromedial temporal horn). When we compared these AFIDs visually (**Figure S2**), we could appreciate that subcortical AFIDs closer to the midline were comparable, but more peripheral AFIDs had lower AFREs using fMRIPrep. Since the inception and data collection of this work, updates have been released for both Lead-DBS and fMRIPrep. To ensure the continued validity of our findings, we compared the registration errors between aforementioned older versions and updated counterparts (Lead-DBS v3.1.0 and fMRIPrep v21.0.1). **Figure S3** demonstrates that overall, most AFREs were similar between software versions. The updated version of Lead-DBS had improved AFREs for AFID02 (PC) and AFID04 (PMJ). fMRIPrep had improved AFREs for AFID02 (PC), AFID04 (PMJ), AFID14 (PG), and AFID 28 (left indusium griseum), and a worse AFRE for AFID25 (right inferior anteromedial temporal horn).

### 3.4 DBS Electrode Tip Position

We analyzed AFREs at various anatomical landmarks; however, in the context of DBS studies, registration error should ideally be measured at the location of the electrode. We first sought to determine if the AFRE acquired at AC and PC could explain electrode position variation in stereotactic space. **Figure 4a** shows the distribution of electrode tips in MNI space with a mesh of the STN superimposed to provide anatomical context (Ewert et al., 2018). As a measure of electrode tip variance, we calculated each subject’s displacement from the mean electrode tip position in all axes and the Euclidean displacement (**Figure 4a)**. The median (IQR) Euclidean displacement was 2.50 mm (1.74-3.33 mm) for the right electrode, and 2.46 mm (1.74-3.44 mm) for the left electrode. Variance of electrode position explained by AFRE at AC and PC is summarized in **Table 1**. A significant amount of the electrode tip displacement was explained by AFREs at both AC and PC. Electrode tip displacement in the y and z axes was especially well explained by AFRE at PC, with 19% and 17% of the variance in the y axis explained, and 17% and 18% of the variance in the z axis explained for the left and right electrode tips respectively (**Figure 4b**). These same patterns remained when using a Spearman’s rank correlation (**Figure S3**). Electrode tip displacement could not be explained by other demographic variables (**Figure S3** and **Table S5**). Electrode displacement was not correlated with age or disease duration in any axis (Spearman’s rank correlation) and was not significantly different between sex, rater pair, modality, side, or implant order (Wilcoxon rank sum test). Finally, including all variables with AFRE in a multivariate linear regression did not change the results (**Table S4**).

**Figure 4.**
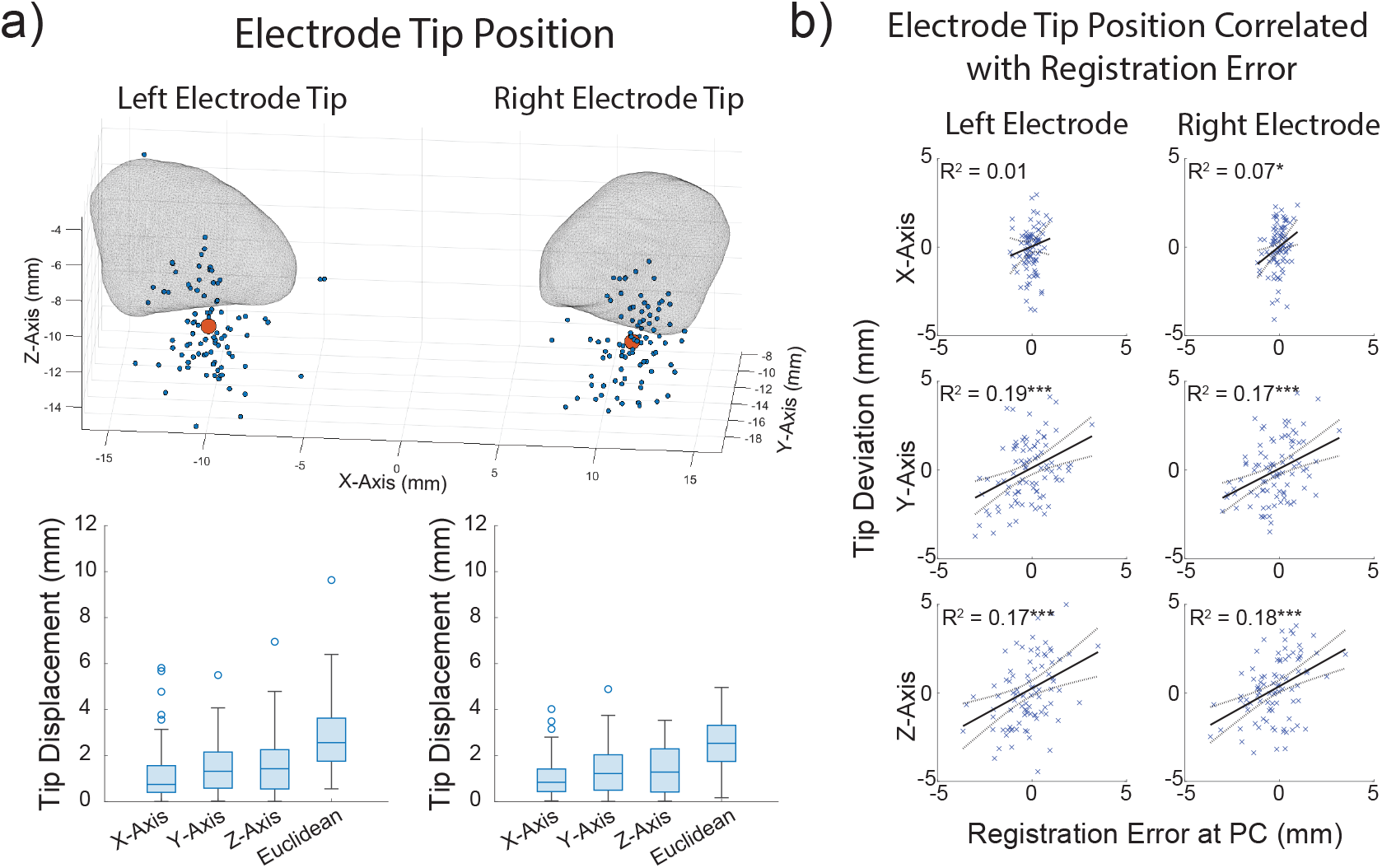
Electrode position in template space is correlated with registration error. (a) 3D scatterplot of all deep brain stimulation (DBS) electrode tip positions (blue, n = 89) and the mean electrode tip position (red) in MNI space, with a mesh of the subthalamic nuclei (Ewert et al., 2018) overlayed (top). Boxplots of each electrode tip’s displacement from the mean electrode tip position in all axes. (b) Correlating left and right electrode tip position with registration error at the posterior commissure (PC) in all axes, (Simple linear regression (n=86), * p<0.05, *** p<0.001). There was a significant correlation between AFRE at PC in the y and z axes and electrode tip displacement in those axes (p<0.001). See Table S6 for more details.

### 3.5 Subcortical AFIDs Correlated with Electrode Position

We next explored whether AFREs at different AFIDs explained the variance of electrode tip position. For each AFID, AFREs were correlated with electrode tip displacement in each axis, as previously performed (**Figure 4b**). **Figure 5** shows the variance of electrode tip displacement explained by AFIDs in each axis. Electrode position in the y axis was best explained by registration error for subcortically located AFIDs. Four AFIDs were significantly correlated with electrode tip position after multiple comparison corrections: AFID02 (PC; 1.80 mm, 1.19-2.25 mm), AFID03 (ICS; 1.59 mm, 0.69-2.31 mm), AFID04 (PMJ; 3.28 mm, 2.37-5.19 mm), and AFID14 (PG; 1.80 mm, 0.94-2.57 mm).

**Figure 5.**
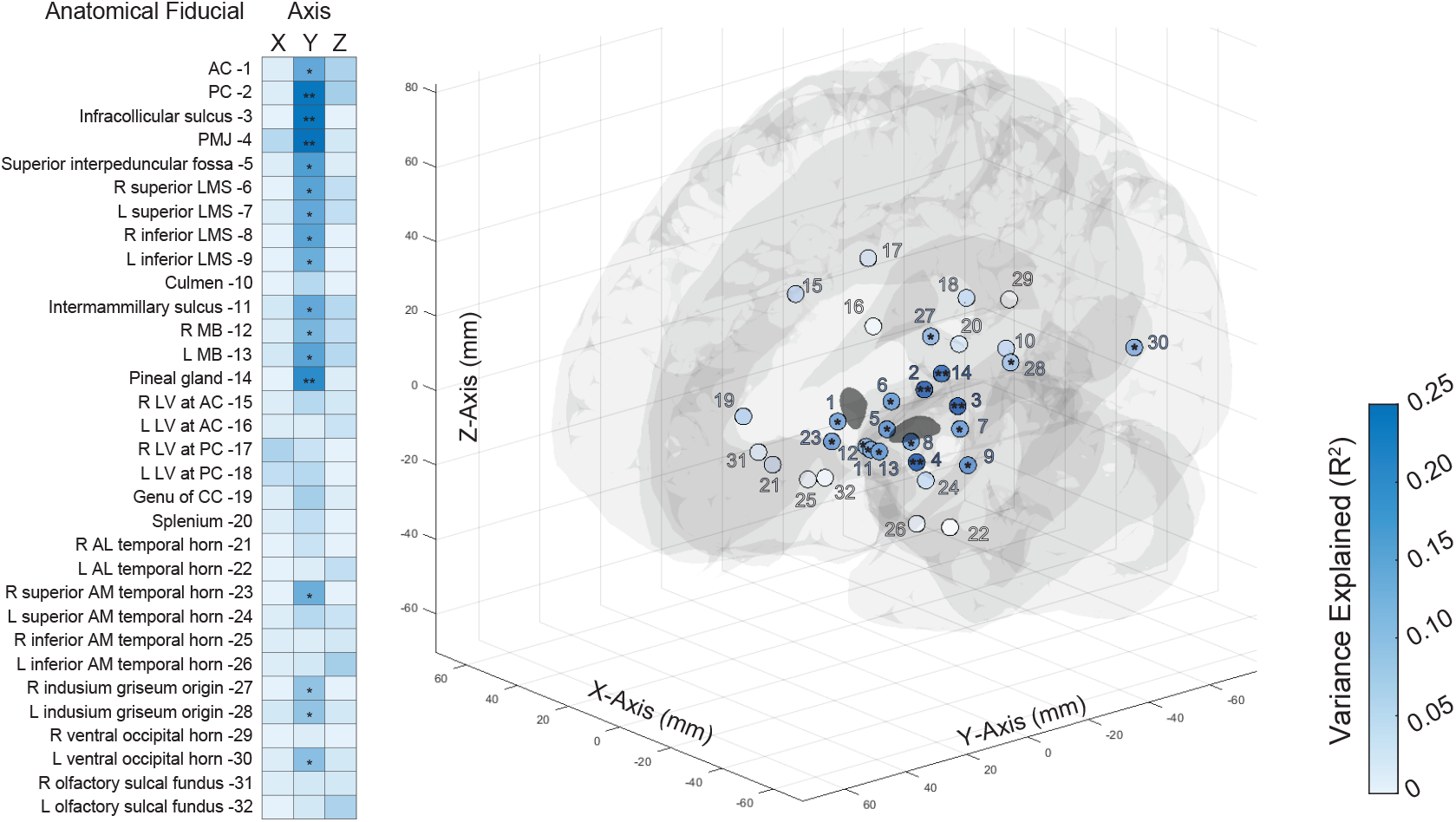
Electrode position in MNI space is correlated with registration errors of anatomical fiducials close to the subthalamic nucleus. Heatmap of electrode tip position variance explained (R^2^) by anatomical fiducial registration error, (Simple linear regression (n=48), * p<0.05, ** p<0.05/32; left) and 3D scatterplot of 32 AFIDs color mapped by R^2^ in the y axis with significance indicated (right). Subcortical AFIDs in close proximity to the electrode target (subthalamic nucleus) had AFREs more correlated with electrode displacement. Specifically, four AFIDs had AFREs significantly correlated (p<0.05/32) with electrode tip displacement in the y axis: AFIDs 02 (posteri- or commissure, PC), 03 (infracollicular sulcus, ICS), 04 (pontomedullary junction, PMJ), and 14 (pineal gland, PG).

To further understand these correlations, we analyzed AFREs at the identified locations using PCA. **Figure 6a** illustrates these AFREs and tip positions as 3D point clouds in MNI space. AFREs at AFID02, AFID03, and AFID14 had a similar 3D distribution, with almost identically oriented PrC axes (**Table S6**). Furthermore, AFREs at these AFIDs were strongly correlated along their PrCs (**Figure 6b**), with the variance explained ranging between 48% to 85% (p<0.001 for all correlations across PrCs). AFID04 had a different AFRE distribution, with a greater contribution from the Cartesian z axis to its first PrC (**Table S6**). The AFRE for AFID04 was less correlated with AFREs for AFID02, AFID03, and AFID14 (**Figure 6b**). Given this strong covariance of AFREs, we performed PCA across all axes of these four AFIDs (i.e. 12 features), and the top 4 PrCs explained 89% of the total variance. The first PrC largely weighed the y axes of AFID02, AFID03, and AFID14, and a combination of the y and z axes of AFID04. The second PrC placed a greater weight on the z axis of AFID04, suggesting the presence of a component of registration error unique to this AFID.

**Figure 6.**
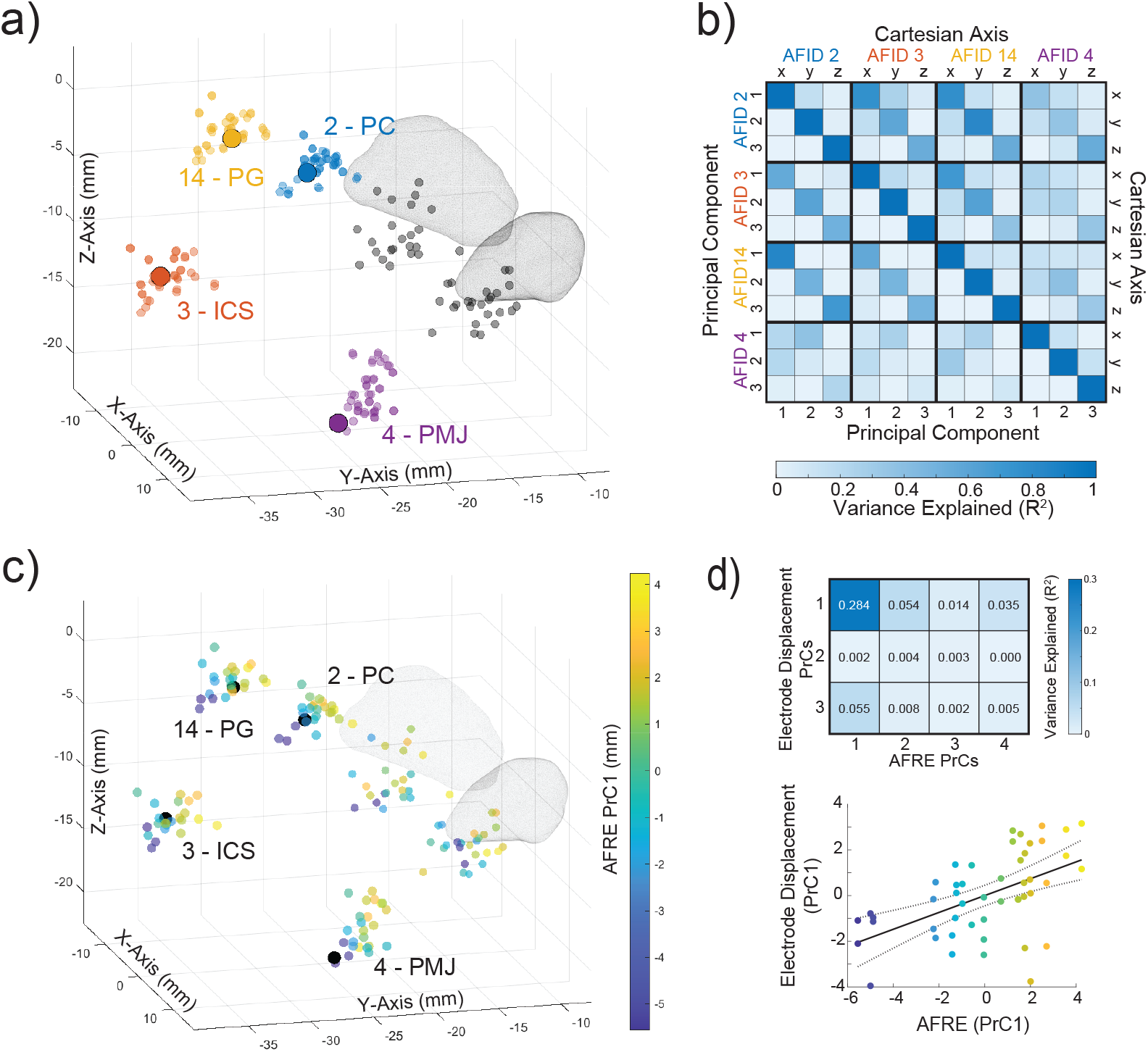
Registration errors covary across a set of subcortical anatomical fiducials close to the DBS target and explain the largest component of variance in electrode position. (a) 3D scatter plot in MNI space with each subject’s transformed anatomical fiducial placements: AFID 02 (posterior commissure, PC), 03 (infracollicular sulcus, ICS), 04 (pontomedullary junction, PMJ), and 14 (pineal gland, PG). The large solid points represent the consensus placements in MNI space. The black transparent points represent the transformed placements of each subject’s left and right electrode tips, with a mesh of the subthalamic nucleus (Ewert et al., 2018). (b) Correlation matrix of AFREs across four AFIDs (left), shown in both standard Cartesian coordinates (top right) and along each AFID’s PrCs (bottom right). (c) PCA across all AFID axes (12 features) was performed, and AFRE was projected on the first PrC. Each subject’s AFRE along the first PrC was linearly mapped to a Parula colour map and projected on a 3D scatter plot in MNI space. (d) Correlation matrix of the top four AFRE PrCs with the electrode tip displacement projected onto its PrCs (top) and scatterplot of AFID AFREs and electrode tip displacement along their first PrCs (bottom).

Finally, we sought to determine if these observed patterns of AFREs across independently placed AFIDs explained the variance observed in electrode position. The first PrC of AFREs is illustrated in **Figure 6c**, which colour maps each subject’s AFRE along this PrC. Next, we performed PCA on the electrode tip displacements, demonstrating a 3D distribution similar to AFID02, AFID03, and AFID14 (**Table S6**). We correlated electrode position along its PrCs to AFREs along their top four PrCs (**Figure 6d**). The first PrC of electrode position was significantly correlated with the first PrC of AFREs, with 28.4% of the electrode tip variance along the first PrC explained (p<0.001). Across all 48 electrodes, this corresponds to a median (range) of 0.64 mm (0.02-2.05 mm) of error in electrode position that can be explained by AFRE.

## 4 Discussion

In this study, we used AFIDs to quantify localization and registration accuracy, and used computed vectors to explain variance in the localization of DBS electrodes. We first demonstrated that DBS electrodes, AC, and PC could be accurately and reliably localized on clinical images (0.63-0.74 mm). AFREs across AFIDs ranged from 1.49 mm to 6.85 mm, and subcortical AFIDs close to the midline had the lowest AFREs. In contrast, DBS electrode positions in stereotactic space had median Euclidean displacements of 2.53 mm and 2.56 mm from the right and left mean tip positions respectively. Finally, we explored the effects of AFRE on electrode position in stereotactic space. AFREs of AFIDs close to the DBS target covaried with each other and were strongly correlated with electrode position, suggesting that common spatial patterns of misregistration to stereotactic space can be detected and accounted for.

### 4.1 Inter-Rater Localization Distance (LD)

Accurate localization of electrode contacts is essential to study the effects of DBS. Despite this, to our knowledge there was only one study that has investigated the reliability of contact localization (Lofredi et al., 2022). We replicate these results (LD ranging from 0.52 mm to 0.75 mm) using the Lead-DBS pipeline, obtaining a median Euclidean LD of 0.73 mm and 0.74 mm for the right and left electrodes respectively. The slightly higher error may be a consequence of the current study using mostly post-operative MRIs to localize DBS electrodes (78.7%), which featured significantly higher LDs when compared to CTs. This is in-line with Lofredi et al. 2022, which found inter-rater LD on post-operative CT to be statistically lower in the x- and y-axes when compared to post-operative MRI. Additionally, we directly compared electrode LDs to LDs obtained at AC (0.68 mm) and PC (0.63 mm). Overall, these results demonstrate that DBS electrode contacts can be as accurately localized as routinely used stereotactic landmarks, roughly within the scale of a voxel on clinical imaging.

### 4.2 Registration Accuracy

Beyond electrode localization, accurate registration of individual images to stereotactic space is a common step in neuroimaging studies investigating group or population-level effects (Barow et al., 2014; Horn et al., 2017; Jeon et al., 2022). This can be particularly important in studying DBS, where clinical effects depend on millimetric accuracy (Kremer et al., 2023; Zhang et al., 2021). To this end, there has been important work to optimize the accuracy of registering individual images to stereotactic space, particularly in subcortical structures (Avants et al., 2010; Ewert et al., 2019; Schönecker et al., 2009). Measures of accuracy have typically relied on voxel overlap measures, like the Jaccard similarity and Dice kappa coefficients (Ewert et al., 2019; V. S. Fonov et al., 2009; Rohlfing, 2012; Vogel et al., 2020). Fiducial based metrics, such as AFRE used in the current study, have rarely been investigated in the context of stereotactic registration, but have the advantage of providing millimetric estimates of registration accuracy. We previously demonstrated that individual fiducials can be quickly placed (of which AC and PC are generally already identified for DBS procedures by the surgical team) and can capture misregistration not observed with voxel overlap metrics (Lau et al., 2019; Miller et al., 2023). Motivated to quantify AFRE in a population of patients who underwent STN DBS for PD, we used our manual placements of each subject’s AC and PC obtaining median AFREs of 1.39 mm and 1.42 mm at AC and PC respectively. This was significantly higher than the LDs at these locations, on an order of two to three times the LD. Therefore, AFRE adds a substantial degree of variance compared to localization error, consistent with previous work (Abbass et al., 2022). We then expanded our analysis to investigate 32 previously identified AFIDs, demonstrating AFREs ranging from 1.49 mm to 6.85 mm with subcortical AFIDs at the midline having the lowest AFREs, which are closest to the conventional targets for DBS.

It is difficult to compare the AFREs we obtained given the limited reports of fiducial-based metrics for evaluating registration from subject to stereotactic space. The initial study describing the AFIDs framework found AFREs ranging from 0.36 mm to 4.51 mm (Lau et al., 2019). The higher AFREs obtained in the current study was expected given the use of clinical images. We subsequently explored AFREs in a set of patients undergoing DBS for PD using non-linear registration (Abbass et al., 2022); a subset of these patients who were also included in the current study, giving us an opportunity to provide proof-of-concept comparison of two commonly used open-source pipelines for neuroimaging analyses: fMRIPrep and Lead-DBS (see **Table S1** for details). Key differences between these pipelines include additional nonlinear SyN registrations that consecutively focused on optimization of registration of subcortical areas, a process termed “subcortical refinement” in Lead-DBS (Ewert et al., 2019). We found that the majority of midline and subcortical AFIDs had similar AFREs using both pipelines, but the Lead-DBS pipeline had decreased AFREs for midline AFIDs located more posteriorly (the pineal gland and splenium). This difference was presumably due to the additional subcortical refinement in Lead-DBS, which aims to provide more accurate registration in anatomically relevant regions for DBS (Ewert et al., 2019). Of note, this comparison was not meant to be exhaustive but rather to be used as an example to demonstrate the utility of the AFIDs framework for providing focal (millimetric) estimates of differences between software versions and packages.

There are limited reports of other groups providing fiducial-based metrics for registration accuracy. Schönecker et al. (2009) computed fiducial-based metrics for their novel registration algorithm using a linear three-step registration focusing on the basal ganglia. The authors computed the root-mean-square error (RMSE) across Cartesian axes for 16 fiducials, rather than the Euclidean-based metric we report. This group reported RMSEs of 1.26 ± 0.70 mm at AC and 0.93 ± 0.72 mm at PC. To directly compare with this prior work, we recomputed our measures as RMSE and obtained a similar RMSE at PC (0.92 ± 0.49 mm), and a lower RMSE at AC (0.97 ± 0.52 mm; **Table S4**). Horn et al., (2017) transformed rater placed AC and PC coordinates from stereotactic space to individual images and calculated the RMSE in each axis. Across AC and PC, RMSE was 0.29 mm (x axis), 1.59 mm (y axis), and 1.16 mm (z axis). In contrast, we obtained higher errors in the x axis (0.42 mm) and z axis (1.40 mm), and lower errors in the y axis (1.22 mm; **Table S4**). Overall, the registration accuracy we obtained in this study is in keeping with previous reports. One advantage of the AFIDs framework is that it leverages open resources and tools, developed with full transparency in mind so that others may freely use, adopt, and modify.

### 4.3 Registration Errors Covary with Electrode Location

The true electrode contact location for a given DBS patient in stereotactic space is unknown. Previous studies have relied on manually obtained fiducial localizations or ROI segmentations near the electrodes, indirectly measuring the effect of misregistration (Ewert et al., 2019; Lau et al., 2019; Vogel et al., 2020). The assumption that nearby anatomical structures share misregistration is inherent to this work; however, to our knowledge, the relationship of misregistration between anatomically distinct regions has not been investigated. AFRE provides a 3D metric with both magnitude and direction which can be leveraged to detect systematic patterns of misregistration between distinct AFIDs. In this study, we find that AFIDs close to each other share similar AFREs. Specifically, we observed that AFID02 (PC), AFID03 (ICS), and AFID14 (PG) had highly correlated AFREs (**Figure 5c**). In fact, these AFREs varied along almost identical independent axes and shared a significant amount of variance. This suggests that misregistration can be similar (in magnitude and direction) across anatomically distinct regions, adding to the validity of previous studies measuring registration accuracy.

Since AFREs across AFIDs can covary, systematic spatial patterns of misregistration may explain some variance in DBS electrode position. We found that our electrode tips had median (IQR) Euclidean displacements 2.50 mm (1.74-3.33 mm) and 2.46 mm (1.74-3.44 mm) for the right and left electrodes respectively. Exploring all 32 AFIDs, we found that the AFIDs closest to the DBS target of the study population (STN) explained a significant amount of variance in electrode tip position. Additionally, electrode position and AFREs at AFID02 (PC), AFID03 (ICS), and AFID14 (PG) all varied along similar independent axes. This suggested that common patterns of misregistration between these AFIDs may have influenced electrode positions. To further support this, we correlated electrode positions along their PrCs with AFREs projected onto their PrCs. We found that the first PrC of AFREs explained 28.4% of the variance in electrode tip position, representing 0.64 mm (0.02-2.05 mm). The patterns of misregistration depend on specific pipeline parameters and software versions, the impact of which can be evaluated more systematically with this framework.

### 4.4 Practical and Clinical Implications

Our findings have important implications for studies that answer population level questions by registering individual brain scans to a stereotactic space. This work is especially relevant for DBS and other stereotactic applications where results are pooled across multiple sites and studies, with clinical effects that depend on millimetric accuracy (Li et al., 2016). As previously discussed, variance in electrode position can be influenced by many factors including application accuracy, LD, and AFRE. Without dissociating these factors, our contacts had a median Euclidean displacement of 2.47 mm, which can be used to contextualize the LDs and AFREs we report. On average, LD of the contacts adds ~0.75 mm of uncertainty to the contact position. Since AFRE cannot be known at the contact location, we can only speculate about the uncertainty added by misregistration. Our most accurate AFREs were in the ~1.50 mm range, and this is likely a conservative estimate of the uncertainty in contact position specifically added by registration error. Some of this registration error (up to 2.05 mm) could be explained by regional AFIDs close to the DBS target location. Furthermore, the extent to which these multiple sources of error accrue remains poorly understood and the framework presented provides a means to uncouple the different components.

Ultimately, the main goal for most of these studies is to explain clinical outcomes by the spatial variance of the volume of tissue activated (VTA) related to application accuracy, providing clinicians with ideal targets (Barow et al., 2014; Horn, 2019; Horn et al., 2017; Zhang et al., 2021). Considering these effects on VTA analyses requires an understanding of typical DBS electrode VTAs, which have been reported to range from 30 to 116 mm^3^ (Chen et al., 2022; Maks et al., 2009). Assuming a spherical VTA of 100 mm^3^ (radius 2.89 mm), the Dice coefficients are: 0.74 for a 1 mm shift, 0.50 for a 2 mm shift, and 0.29 for a 3 mm shift. Therefore, the LDs and AFREs we observe have the potential to change more than half of the VTA in stereotactic space. Ideally, these errors are random, without any bias, as we observed with many AFREs such as with AFID02, AFID03, and AFID14 (**Figure 6a**), in which case this added variance may be overcome with sufficiently powered studies. Our approach using detected patterns of misregistration across AFIDs to explain variance in contact position may result in more optimal estimates of target locations.

### 4.5 Limitations and Future Directions

Although we can measure AFREs at validated AFIDs and correlate this with DBS electrode position, we are unable to determine if registration errors are biased at the electrode position since the ‘true’ electrode position in stereotactic space is unknown. This issue may partially be addressed by identifying and validating an anatomical landmark at or closer to the target, which can be challenging since in this case the target (STN) can be difficult to identify on the commonly acquired clinical scans. Another limitation is our use of gadolinium-enhanced images, which have not been optimized by automated registration pipelines and thus may have higher registration errors than images without gadolinium. Despite these potential concerns about gadolinium, the AFREs we obtained are in keeping with previous work reporting registration errors (Horn et al., 2017; Schönecker et al., 2009). Furthermore, gadolinium-enhanced scans typically represent the main reference image used during stereotactic planning due to the combination of high-quality anatomical imaging, and the ability to visualize cerebral vasculature without the need for additional multimodal fusion for trajectory optimization.

Previous work has found that the addition of multispectral data (specifically T2w scans) improve the registration process (Ewert et al., 2019). In this first study on AFIDs for DBS applications, we opted to avoid introducing the complexity of additional multimodal data (T2w images) and focussed on registration between the reference anatomical (T1w) scan and MNI space. The AFIDs framework can still be applied in a multi-spectral context. More specifically, we can evaluate the co-registration of multimodal scans at the individual subject level. Then, we can evaluate registration of a population using AFIDs on a subject’s “base” scan. Future studies aim to explore the impact of multi-spectral registration on electrode localization variance in template space.

AFIDs require manual placement, which takes expertise. One barrier to adoption is that placement has been perceived to be overly time-consuming, although in our experience trained raters can complete the protocol within 15-20 minutes. This motivated the open release of a curated set of manually placed AFIDs on 14 templates and 132 individual scans, including the data used in this study (Taha et al., 2023). Since we have shown that only a few AFIDs are necessary to explain variations in electrode position for STN DBS, it may be determined that only a subset may be necessary for a given indication. In fact, AC and PC are commonly already manually defined when planning a DBS procedure and can be quickly leveraged to calculate AFREs and explain electrode position variance. Future tools can furthermore benefit from automatic placement of AFIDs, which can be easily incorporated into established open toolboxes for neuroimaging (Esteban et al., 2019; Horn & Kühn, 2015).

We compared registration accuracy in two commonly used open tools for neuroimaging analysis: Lead-DBS and fMRIPrep. We wish to emphasize that this comparison was not meant to be formal or exhaustive but to be used as a proof-of-concept of how the AFIDs framework can be employed to evaluate registration accuracy across different tools and versions. We selected Lead-DBS given it is the most widely adopted tool for DBS electrode localization, its transparent and open development, and given the clinical context of our dataset. We furthermore selected fMRIPrep based on previous experience (Abbass et al., 2022) and given the direct development of this application from the open Brain Imaging Data Structure (BIDS) standard (Gorgolewski et al., 2016). Both Lead-DBS and fMRIPrep employ ANTS for registration on the back end (Avants et al., 2008), and have active user and support communities. Future studies can use this framework for more detailed quality control, software testing with continuous integration, optimization and comparison of different software versions and pipelines, as well as different registration algorithms.

### 4.6 Concluding Remarks

In summary, we used the AFIDs framework to investigate localization and registration accuracy, as well as explain variance in electrode position related to transformations to stereotactic space. The AFIDs framework is an open resource, and curated AFID placements for various imaging datasets have been released (Taha et al., 2023). Using this framework, we provide AFREs at different anatomical locations to estimate the magnitude and direction of registration accuracy. To our knowledge, these represent the first millimetric estimates of registration accuracy in DBS, allowing uncoupling of registration-related factors from other sources of variance in electrode position. Additionally, we show that AFREs can covary, identifying potential systematic spatial patterns of misregistration in a dataset, that can explain a significant portion of the variance observed in electrode positions. Accounting for these registration errors has the potential to dissociate application error from registration error in a stereotactic space, improving the spatial specificity of group-level analyses.

## Data Availability

All data and scripts used are available online at: https://github.com/afids/afids-dbs

## Data and Code Availability

All raw and processed data along with processing scripts used for data analysis (MATLAB 2022a) used in this manuscript are available at https://github.com/afids/afids-dbs.

## Author Contributions

M.A. contributed to conceptualization, data curation, formal analysis, methodology, and writing. A.T. contributed to conceptualization, methodology, and writing. G.G., B.S., and A.C. contributed to conceptualization, data curation, and writing. M.J., K.M. A.P., and T.P. contributed to conceptualization and writing. J.L. contributed to conceptualization, methodology, supervision, and writing.

