## Supplemental Materials for "The impact of localization and registration accuracy on estimates of deep brain stimulation electrode position in stereotactic space"

**Table S1 – Registration Parameters.**

| Lead-DBS version 2.3.2* | | | |
|---|---|---|---|
| **Parameter** | **Rigid** | **Affine** | **SyN (x3)** |
| *Gradient Step* | 0.25 | 0.15 | 0.3, 4, 3 |
| *Metric* | MI | MI | MI |
| *Metric Parameters* | 1.25, 32, Random, 0.25 | 1.25, 32, Random, 0.25 | 1.25, 32, Random, 0.25 |
| *Convergence* | 1000 x 500 x 250 x 0, $10^{-7}$, 10 | 1000 x 500 x 250 x 0, $10^{-7}$, 10 | 1000 x 500 x 500 x 0, $10^{-7}$, 7 |
| *Shrink Factors* | 12 x 8 x 4 x 1 | 12 x 8 x 4 x 1 | 8 x 4 x 4 x 1 |
| *Smooth Sigmas* | 5 x 4 x 3 x 1 | 5 x 4 x 3 x 1 | 4 x 3 x 1 x 1 |
| **fMRIPrep version 1.5.4** | | | |
| **Parameter** | **Rigid** | **Affine** | **SyN** |
| *Gradient Step* | 0.1 | 0.1 | 0.1, 3, 0 |
| *Metric* | MI | MI | MI |
| *Metric Parameters* | 1, 32, Regular, 0.25 | 1, 32, Regular, 0.25 | 1, 32 |
| *Convergence* | 1000 x 500 x 250 x 0, $10^{-6}$, 10 | 1000 x 500 x 250 x 0, $10^{-6}$, 10 | 100 x 100 x 70 x 50 x 0, $10^{-6}$, 10 |
| *Shrink Factors* | 12 x 8 x 4 x 2 | 12 x 8 x 4 x 2 | 10 x 6 x 4 x 2 x 1 |
| *Smooth Sigmas* | 4 x 3 x 2 x 1 | 4 x 3 x 2 x 1 | 5 x 3 x 2 x 1 x 0 |

Registration parameters implemented in the current study using Lead-DBS (v.2.3.2; netstim.gitbook.io/leaddbs) and in our previous study (Abbass et al., 2022) using fMRIPrep (v1.5.4). fMRIPrep (v.21.0.1) and Lead-DBS (v.3.1.0) was also run with default parameters for comparisons and to account for more recent developments. For detailed documentation and visualizations, we refer readers to LeadDBS (https://netstim.gitbook.io/leaddbs/lead-dbs/) and fMRIPrep (fmriprep.org/en/1.5.4/) official documentation.

*Lead-DBS SyN registration was completed in three stages, a whole brain stage followed by two stages focusing on subcortical areas.

**Table S2 – Demographic data for human raters**

| Rater | Description |
|---|---|
| A | Neurophysiologist with 10+ years of experience in DBS, neuroimaging software, and neuroanatomy |
| B,C,D | Neurosurgical residents with 5+ years of experience in neuroanatomy and neuroimaging software |

Demographic information for the four raters who localized electrodes and placed anterior and posterior commissure coordinates in this current study, with relevant neuroanatomical experience.

**Table S3 – Study Demographics.**

| | | Subjects with AC/PC (n=89) | Subjects with 32 AFIDs (n=24) |
|---|---|---|---|
| **Age (years)** | *Mean* | 60.5 | 60.1 |
| | *Std* | 6.1 | 5.5 |
| **Disease Duration (years)** | *Mean* | 11.01 | 11.71 |
| | *Std* | 4.21 | 3.68 |
| **Sex** | *Male* | 61 (68.5%) | 14 (58.3%) |
| | *Female* | 28 (31.5%) | 10 (45.8%) |
| **Implanted first** | *Right* | 69 (77.5%) | 13 (54.2%) |
| | *Left* | 20 (22.5%) | 11 (45.8%) |
| **Post-operative Imaging** | *CT* | 19 (21.3%) | 4 (16.7%) |
| | *MRI* | 70 (78.7%) | 20 (83.3%) |
| **Rater Pair** | *Pair 1 (MA/BS)* | 43 (48.3%) | 11 (45.8%) |
| | *Pair 2 (GG/AC)* | 46 (51.7%) | 13 (54.2%) |

Demographic information of all 89 subjects who underwent the Lead-DBS protocol by two raters with manual placements of the anterior commissure (AC) and posterior commissure (PC). Demographic information of a subset of 24 subjects with manual placements of 32 anatomical fiducials by five raters is also provided.

**Table S4 – Summary of localization distance metrics.**

| | | Localization Distance (mm) | | | | | | |
|---|---|---|---|---|---|---|---|---|
| | | **Min** | **LQ** | **Median** | **UQ** | **Max** | **Mean** | **Std** |
| **Right Electrode** | *X Axis* | 0.01 | 0.11 | 0.27 | 0.41 | 1.03 | 0.30 | 0.05 |
| | *Y Axis* | 0.00 | 0.12 | 0.26 | 0.44 | 1.55 | 0.33 | 0.10 |
| | *Z Axis* | 0.00 | 0.32 | 0.50 | 0.76 | 2.54 | 0.57 | 0.19 |
| | *Euclidean* | 0.05 | 0.53 | 0.73 | 1.06 | 2.58 | 0.82 | 0.19 |
| **Left Electrode** | *X Axis* | 0.00 | 0.13 | 0.25 | 0.44 | 1.10 | 0.31 | 0.06 |
| | *Y Axis* | 0.00 | 0.13 | 0.26 | 0.57 | 1.56 | 0.39 | 0.12 |
| | *Z Axis* | 0.00 | 0.24 | 0.44 | 0.73 | 1.55 | 0.52 | 0.14 |
| | *Euclidean* | 0.12 | 0.50 | 0.74 | 1.10 | 1.77 | 0.82 | 0.17 |
| **Anterior Commissure** | *X Axis* | 0.00 | 0.11 | 0.18 | 0.33 | 0.88 | 0.24 | 0.03 |
| | *Y Axis* | 0.00 | 0.12 | 0.28 | 0.53 | 2.38 | 0.38 | 0.14 |
| | *Z Axis* | 0.00 | 0.16 | 0.51 | 0.81 | 3.63 | 0.57 | 0.29 |
| | *Euclidean* | 0.00 | 0.48 | 0.68 | 1.03 | 3.70 | 0.81 | 0.32 |
| **Posterior Commissure** | *X Axis* | 0.00 | 0.09 | 0.20 | 0.36 | 1.03 | 0.25 | 0.04 |
| | *Y Axis* | 0.00 | 0.13 | 0.27 | 0.52 | 2.12 | 0.36 | 0.12 |
| | *Z Axis* | 0.00 | 0.20 | 0.39 | 0.68 | 2.51 | 0.49 | 0.19 |
| | *Euclidean* | 0.00 | 0.41 | 0.63 | 1.01 | 2.66 | 0.75 | 0.21 |

Summary metrics of localization distances obtained between raters for deep brain stimulation electrodes, anterior commissure (AC) and posterior commissure (PC), n=89. Absolute distances in each axis and the Euclidean Distance are reported in millimeters. For each, the minimum (min), lower quartile (LQ), median, upper quartile (UQ), maximum (max), mean and standard deviation (Std) is reported.

**Table S5 – Summary of registration error metrics for AC and PC.**

| | | Registration Error (mm) | | | | | | | |
|---|---|---|---|---|---|---|---|---|---|
| **Current Study** | | **Min** | **LQ** | **Median** | **UQ** | **Max** | **Mean** | **Std** | **RMSE[b]** |
| *Anterior Commissure* | *X Axis* | 0.03 | 0.14 | 0.26 | 0.41 | 1.35 | 0.30 | 0.22 | 0.40 |
| | *Y Axis* | 0.03 | 0.33 | 0.67 | 1.09 | 3.18 | 0.83 | 0.68 | 1.15 |
| | *Z Axis* | 0.05 | 0.53 | 1.06 | 1.77 | 3.96 | 1.24 | 0.88 | 1.49 |
| | *Euclidean* | 0.33 | 1.05 | 1.39 | 2.38 | 4.38 | 1.70 | 0.90 | - |
| | *RMSE[a]* | 0.18 | 0.57 | 0.79 | 1.35 | 2.53 | 0.96 | 0.52 | - |
| *Posterior Commissure* | *X Axis* | 0.03 | 0.18 | 0.29 | 0.38 | 1.16 | 0.34 | 0.24 | 0.43 |
| | *Y Axis* | 0.14 | 0.41 | 0.77 | 1.28 | 3.17 | 0.97 | 0.72 | 1.23 |
| | *Z Axis* | 0.13 | 0.48 | 0.85 | 1.30 | 3.65 | 1.01 | 0.76 | 1.26 |
| | *Euclidean* | 0.30 | 1.00 | 1.42 | 2.10 | 4.37 | 1.62 | 0.85 | - |
| | *RMSE[a]* | 0.16 | 0.56 | 0.80 | 1.21 | 2.52 | 0.92 | 0.49 | - |
| *AC and PC Combined* | *X Axis* | 0.03 | 0.16 | 0.28 | 0.41 | 1.35 | 0.32 | 0.23 | 0.42 |
| | *Y Axis* | 0.03 | 0.38 | 0.72 | 1.22 | 3.18 | 0.90 | 0.71 | 1.23 |
| | *Z Axis* | 0.05 | 0.51 | 0.97 | 1.43 | 3.96 | 1.13 | 0.82 | 1.40 |
| | *Euclidean* | 0.30 | 1.02 | 1.42 | 2.23 | 4.38 | 1.66 | 0.87 | - |
| | *RMSE[a]* | 0.16 | 0.57 | 0.80 | 1.25 | 2.53 | 0.94 | 0.50 | - |
| **Schönecker et al., 2009** | | | | | | | | | |
| *Anterior Commissure* | *RMSE[a]* | - | - | - | - | - | 1.26 | 0.70 | - |
| *Posterior Commissure* | *RMSE[a]* | - | - | - | - | - | 0.93 | 0.72 | - |
| **Horn et al., 2017** | | | | | | | | | |
| *AC and PC Combined* | *X Axis* | - | - | - | - | - | - | 0.18 | 0.29 |
| | *Y Axis* | - | - | - | - | - | - | 0.97 | 1.59 |
| | *Z Axis* | - | - | - | - | - | - | 0.69 | 1.16 |

Summary metrics of anatomical fiducial registration errors obtained for the anterior commissure (AC), posterior commissure (PC) and their combination, n=89. For each fiducial, absolute registration error in the x, y, and z axes, as well as Euclidean error and root-mean-square error (**RMSE[a]**) across the three axes is reported for comparative purposes. For each, the minimum (min), lower quartile (LQ), median, upper quartile (UQ), maximum (max), mean and standard deviation (Std) is reported. Additionally, RMSE for each of the x, y, and z axes (**RMSE[b]**) was computed for comparison. Previously reported metrics of fiducial-based registration errors (Schönecker et al. 2009 and Horn et al., 2018) are also included.

**Table S6 – Univariate and multivariate analyses used to explain electrode tip displacement in X, Y, and Z axes.**

| | **Univariate Analysis** | | | **Multivariate Analysis** | | |
|---|---|---|---|---|---|---|
| | **Non-Parametric Test (*p*-value)** | | | **T-Statistic (*p*-value)** | | |
| **Independent Variables** | **X-Axis** | **Y-Axis** | **Z-Axis** | **X-Axis** | **Y-Axis** | **Z-Axis** |
| *Age (years)* | (0.116) | (0.234) | (0.758) | 1.33 (0.186) | 1.04 (0.298) | -0.82 (0.412) |
| *Disease Duration (years)* | (0.089) | (0.319) | (0.046) | 1.42 (0.157) | 0.82 (0.414) | -2.44 (0.016) |
| *Sex* | (0.013) | (0.149) | (0.996) | 1.64 (0.104) | -0.22 (0.828) | 0.82 (0.416) |
| *Rater Pair* | (0.822) | (0.804) | (0.972) | 0.20 (0.842) | -0.52 (0.603) | -0.69 (0.494) |
| *Modality (MRI/CT)* | (0.312) | (0.826) | (0.561) | -0.74 (0.46) | 0.19 (0.846) | -0.06 (0.954) |
| *Electrode Side* | (0.348) | (0.967) | (0.527) | -0.83 (0.407) | -0.34 (0.737) | 0.38 (0.708) |
| *Implant Order* | (0.029) | (0.693) | (0.805) | -2.36 (0.020) | -0.82 (0.411) | 0.15 (0.877) |
| *AC Reg Error (mm)* | **(0.001)** | **(<0.001)** | **(<0.001)** | 1.70 (0.092) | 1.85 (0.067) | 2.32 (0.021) |
| *PC Reg Error (mm)* | (0.012) | **(<0.001)** | **(<0.001)** | 0.62 (0.533) | **4.75 (<0.001)** | **3.43 (<0.001)** |
| **Adjusted $R^2$ by multivariate model (*p*-value)** | | | | **0.09 (0.004)** | **0.17 (<0.001)** | **0.19 (<0.001)** |

Univariate and multivariate analysis examining the effect of demographic variables and anatomical fiducial registration errors on electrode location (n=172 electrodes). For Univariate analyses, electrode tip displacement was correlated with continuous variables (age, disease duration, anterior commissure (AC) registration error and posterior commissure (AC) registration error) using a Spearman's rank correlation. Electrode tip displacement was compared between binary variables (sex, rater pair, modality (MRI or CT), electrode side and implantation order) using a Wilcoxon rank sum test. Statistical results for univariate tests are presented, and significant p-values are bolded (alpha = 0.05/9 for each axis). A multivariate linear regression model was used with 9 independent variables. Adjusted $R^2$ values for the whole model and T-statistics for each independent variable is presented. In summary, there results suggest that electrode tip placement is correlated with AC and PC registration errors.

**Table S7 – Summary results of principal components analysis.**

| | | Coefficients | | | |
|---|---|---|---|---|---|
| | | **X-Axis** | **Y-Axis** | **Z-Axis** | **% Explained** |
| **AFID02** | *PrC 1* | -0.10 | 0.96 | 0.25 | 64.0 |
| | *PrC 2* | 0.00 | -0.25 | 0.97 | 32.9 |
| | *PrC 3* | 1.00 | 0.10 | 0.03 | 3.1 |
| **AFID03** | *PrC 1* | -0.15 | 0.97 | 0.21 | 64.4 |
| | *PrC 2* | -0.08 | -0.22 | 0.97 | 31.4 |
| | *PrC 3* | 0.99 | 0.13 | 0.11 | 4.2 |
| **AFID14** | *PrC 1* | -0.13 | 0.97 | 0.21 | 65.0 |
| | *PrC 2* | -0.11 | -0.23 | 0.97 | 30.2 |
| | *PrC 3* | 0.99 | 0.10 | 0.14 | 4.8 |
| **AFID04** | *PrC 1* | 0.00 | 0.33 | 0.94 | 72.9 |
| | *PrC 2* | -0.21 | 0.92 | -0.33 | 23.7 |
| | *PrC 3* | 0.98 | 0.20 | -0.07 | 3.5 |
| **Electrode Displacement** | *PrC 1* | -0.01 | 0.91 | 0.41 | 48.2 |
| | *PrC 2* | -0.27 | -0.40 | 0.88 | 36.5 |
| | *PrC 3* | 0.96 | -0.10 | 0.25 | 15.3 |

| | Coefficients | | | | | | | | | | | | |
|---|---|---|---|---|---|---|---|---|---|---|---|---|---|
| | **AFID 2** | | | **AFID 3** | | | **AFID 14** | | | **AFID 4** | | | |
| | *X* | *Y* | *Z* | *X* | *Y* | *Z* | *X* | *Y* | *Z* | *X* | *Y* | *Z* | *% Explained* |
| **PrC 1** | -0.06 | 0.45 | 0.19 | -0.09 | 0.46 | 0.18 | -0.09 | 0.48 | 0.16 | -0.07 | 0.31 | 0.38 | 46.97 |
| **PrC 2** | 0.01 | -0.23 | 0.36 | 0.01 | -0.34 | 0.35 | -0.01 | -0.30 | 0.33 | 0.07 | -0.02 | 0.61 | 29.40 |
| **PrC 3** | 0.04 | 0.20 | 0.17 | 0.01 | -0.55 | -0.11 | 0.02 | 0.29 | 0.55 | -0.01 | 0.25 | -0.41 | 6.39 |
| **PrC 4** | -0.09 | -0.18 | 0.18 | -0.14 | 0.30 | 0.67 | -0.10 | -0.02 | 0.19 | -0.12 | -0.24 | -0.50 | 5.87 |
| **PrC 5** | -0.10 | -0.32 | -0.06 | -0.10 | -0.01 | 0.16 | -0.04 | -0.18 | -0.21 | -0.15 | 0.86 | -0.11 | 4.28 |
| **PrC 6** | 0.37 | -0.12 | 0.28 | 0.52 | 0.04 | 0.20 | 0.54 | 0.25 | -0.24 | 0.19 | 0.08 | -0.08 | 2.68 |
| **PrC 7** | -0.15 | -0.22 | 0.63 | -0.18 | -0.15 | -0.25 | -0.14 | 0.28 | -0.38 | -0.37 | -0.14 | -0.01 | 1.49 |
| **PrC 8** | 0.00 | 0.48 | -0.21 | -0.08 | -0.47 | 0.47 | 0.04 | 0.03 | -0.44 | -0.26 | -0.07 | 0.05 | 1.14 |
| **PrC 9** | -0.03 | -0.52 | -0.48 | -0.05 | -0.15 | 0.13 | -0.01 | 0.63 | 0.09 | -0.08 | -0.10 | 0.20 | 0.85 |
| **PrC 10** | 0.00 | -0.01 | 0.11 | -0.27 | -0.13 | 0.11 | -0.33 | 0.17 | -0.27 | 0.82 | 0.04 | -0.07 | 0.65 |
| **PrC 11** | -0.11 | 0.01 | -0.01 | 0.74 | -0.04 | 0.06 | -0.65 | 0.02 | -0.05 | -0.06 | 0.01 | -0.02 | 0.22 |
| **PrC 12** | 0.89 | -0.03 | -0.02 | -0.19 | 0.04 | -0.02 | -0.35 | -0.03 | 0.01 | -0.19 | 0.02 | 0.01 | 0.06 |

Principal components analysis performed on individual anatomical fiducials (AFIDs) and electrode tip displacements (top) and across all axes and AFIDs (4 AFIDs x 3 axes, or 12 features; bottom). Coefficients for each principal component (PrC) is presented, along with the percentage of total variance explained by each PrC.

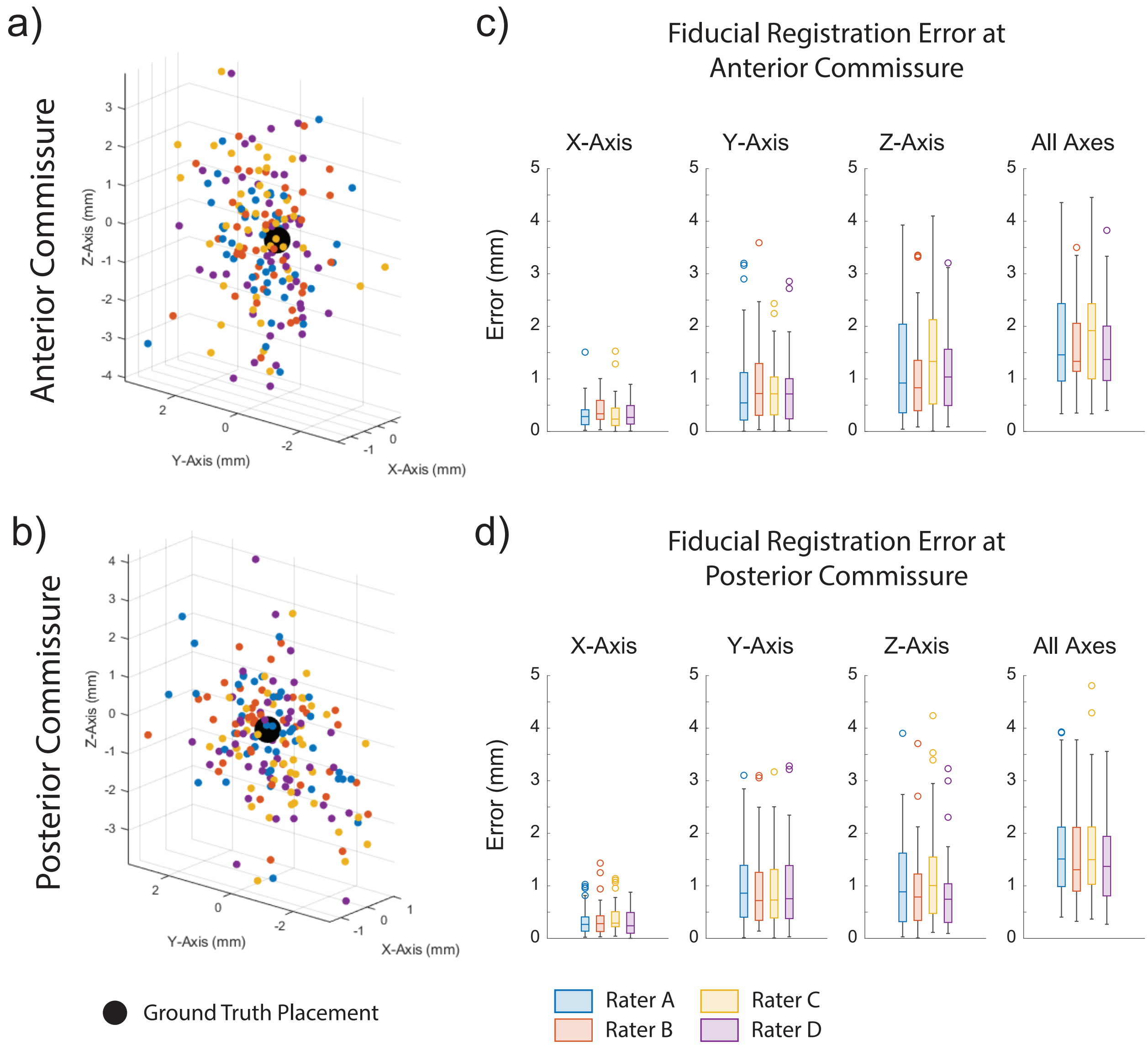


**Figure S1. Anatomical fiducial registration errors are consistent across individual raters.** (a-b) 3D scatterplots of transformed rater placements of anterior commissure (AC) and posterior commissure (PC) and ground truth placement (black). Transformed AC and PC points are color mapped by rater. (c-d) Anatomical fiducial registration errors (AFREs) obtained across individual raters at AC and PC in all axes. There were no significant differences across AFREs independently obtained by individual raters (Wilcoxon rank sum, alpha = 0.05/6 in each axis).

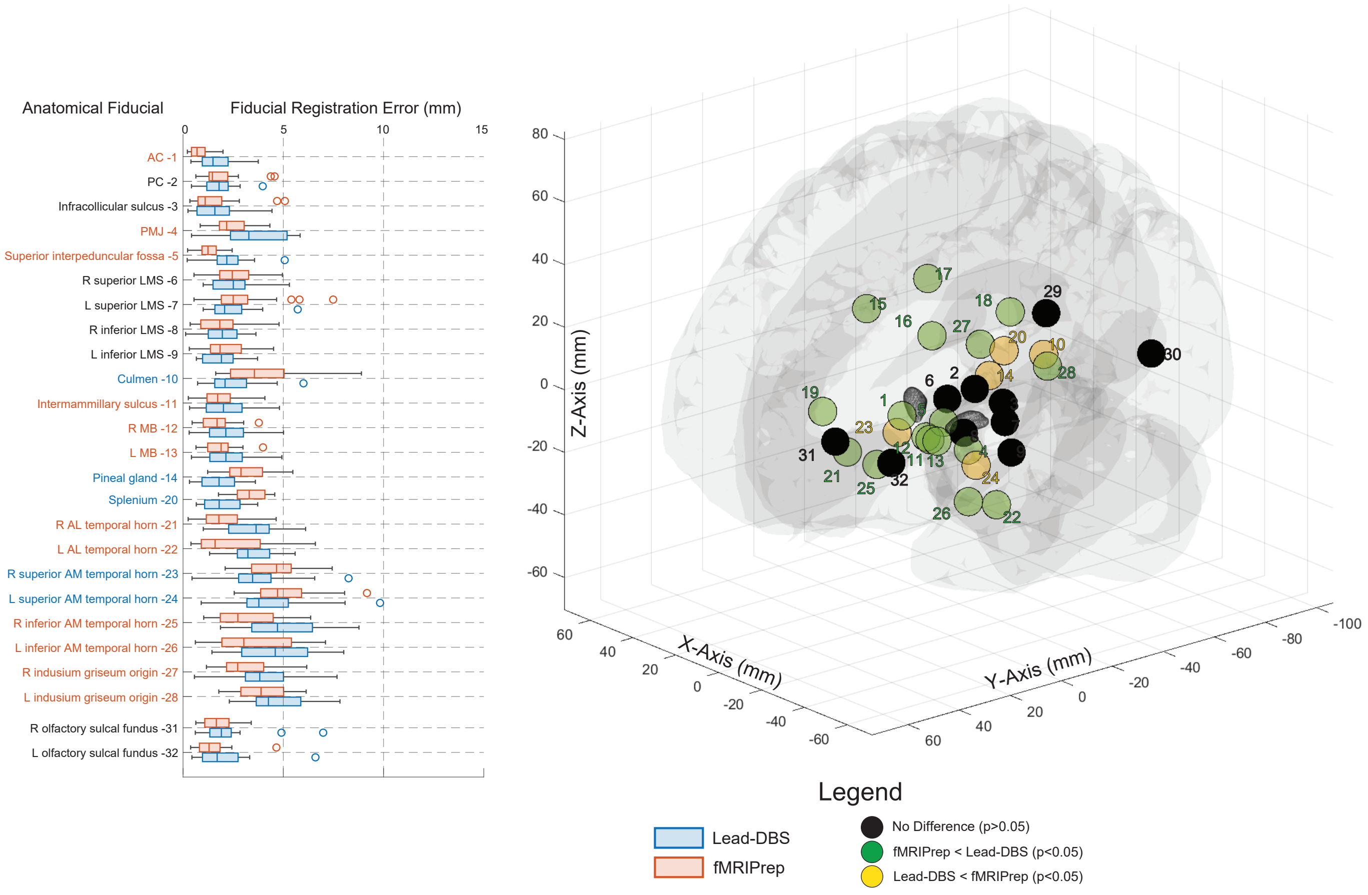


**Figure S2. Anatomical fiducial registration errors compared using Lead-DBS and fMRIPrep toolsets.** Comparing fiducial registration errors (AFREs) obtained across subcortical anatomical fiducials (AFIDs) in the current study using Lead-DBS (v.2.3.2) and in Abbass et al. (2022) using fMRIPrep 1.5.4. Euclidean AFREs obtained by Lead-DBS and fMRIPrep for each AFID are shown using boxplots (left). AFREs are compared using a Wilcoxon signed-rank Test (n=24). AFIDs with a p-value <0.05 are color-coded such that smaller AFREs obtained by Lead-DBS are orange, and smaller AFREs obtained by fMRIPrep are green. Color-mapped AFIDs are plotted in a 3D scatter-plot with an overlying brain mesh and mesh of each subthalamic nucleus (Ewert et al., 2018).

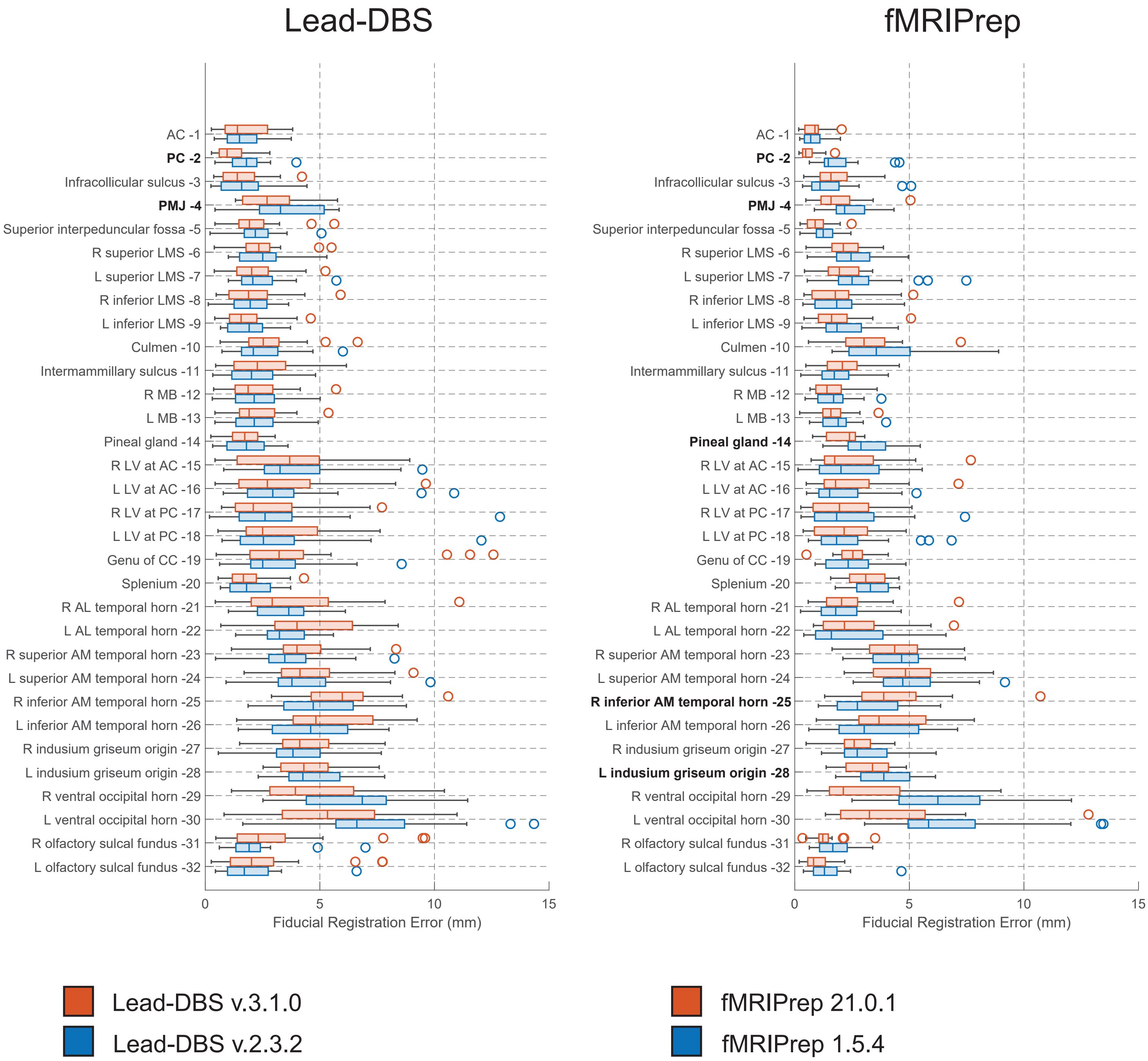


**Figure S3. Anatomical fiducial registration errors compared different versions of Lead-DBS and fMRIPrep toolsets.** Comparing fiducial registration errors (AFREs) obtained across 32 anatomical fiducials (AFIDs) between older versions of Lead-DBS (v.2.3.2) and fMRIPrep (1.5.4) available at the conception of this study, to newer versions available after completion of the manuscript (Lead-DBS v.3.1.0 and fMRIPrep 21.0.1). Euclidean AFREs for each AFID are shown using boxplots (left), with older versions coloured in blue and newer versions colored in red. AFREs are compared using a Wilcoxon signed-rank Test (n=24) with a Bonferroni correction (0.05/32). Significantly different AFIDs are bolded.

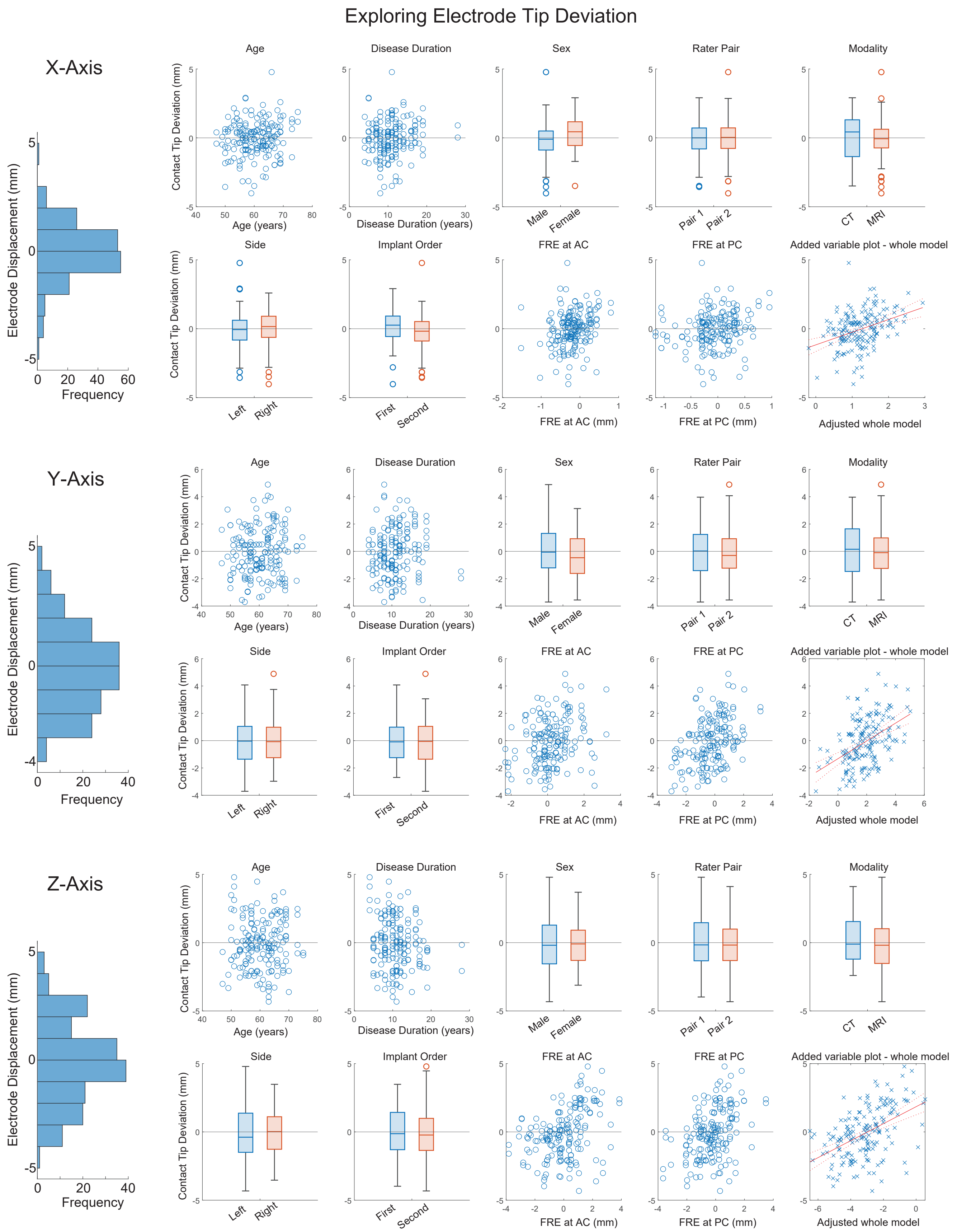


**Figure S4. Electrode tip position in template space is not explained by demographic or imaging variables.** Explaining electrode tip displacement by demographic variables (age, disease duration, sex), rater pair, modality (post-operative CT or MRI used), electrode side, implantation order (first or second side), and anatomical fiducial registration error (AFRE) at the anterior commissure (AC) and posterior commissure (PC). Electrode tip deviation is shown in a histogram for each axis. Electrode tip deviation is plotted against continuous independent variables using a scatterplot and is represented by boxplots for binary independent variables.
